# Onco-Seg: Adapting Promptable Concept Segmentation for Multi-Modal Medical Imaging

**DOI:** 10.64898/2026.01.11.26343874

**Authors:** Ashish Makani, Anjali Agrawal, Anurag Agrawal

## Abstract

Medical image segmentation remains a critical bottleneck in clinical workflows, from diagnostic radiology to radiation oncology treatment planning. We present **Onco-Seg**, a medical imaging adaptation of Meta’s Segment Anything Model 3 (SAM3) that leverages promptable concept segmentation for automated tumor and organ delineation across multiple imaging modalities. Unlike previous SAM adaptations limited to single modalities, Onco-Seg introduces a unified framework supporting CT, MRI, ultrasound, dermoscopy, and endoscopy through modality-specific preprocessing and parameter-efficient fine-tuning with Low-Rank Adaptation (LoRA). We train on 35 datasets comprising over 98,000 cases across 8 imaging modalities using sequential checkpoint chaining on a 4-GPU distributed training infrastructure. We evaluate Onco-Seg on 12 benchmark datasets spanning breast, liver, prostate, lung, skin, and gastrointestinal pathologies, achieving strong performance on breast ultrasound (Dice: 0.752±0.24), polyp segmentation (Dice: 0.714±0.32), and liver CT (Dice: 0.641±0.12). We further propose two clinical deployment patterns: an interactive “sidecar” for diagnostic radiology and a “silent assistant” for automated radiation oncology contouring. We release an open-source napari plugin enabling interactive segmentation with DICOM-RT export for radiation oncology workflows. Code and models are available at https://github.com/inventcures/onco-segment.

## 1. Introduction

Medical image segmentation is fundamental to modern clinical practice. Radiologists rely on precise tumor delineation for diagnosis and treatment response assessment. Radiation oncologists require accurate organ-at-risk (OAR) contouring for treatment planning, where segmentation errors can result in inadequate tumor coverage or excessive normal tissue irradiation [4]. Despite decades of algorithmic development, manual segmentation remains the clinical standard, consuming significant physician time and introducing inter-observer variability [5].

The Segment Anything Model (SAM) series has transformed computer vision by demonstrating that large-scale pretraining enables zero-shot segmentation across diverse domains. SAM1 [1] introduced promptable segmentation with points and boxes. SAM2 [2] extended this to video with memory-based tracking. SAM3 [3], released in November 2025, represents a paradigm shift through Promptable Concept Segmentation (PCS)—the ability to segment all instances of a concept specified by text or image exemplars.

SAM3’s innovations present unprecedented opportunities for medical imaging:

1. **Text-Based Prompting:** Clinicians specify targets using natural language (“liver tumor,” “left parotid gland”) rather than manual annotations.
2. **Multi-Instance Segmentation:** SAM3 identifies all lesions matching a concept—critical for metastatic disease.
3. **Presence Token Architecture:** Decouples recognition from localization, enabling discrimination between similar concepts.
4. **Unified Detector-Tracker Design:** Naturally extends to 3D volumetric data by treating slices as frames.

In this work, we present Onco-Seg, a comprehensive adaptation of SAM3 for medical imaging. Our contributions include: (1) a unified preprocessing pipeline supporting ten imaging modalities; (2) parameter-efficient fine-tuning using LoRA on SAM3’s 848M parameter architecture; (3) training on 35 datasets with over 98,000 cases using sequential checkpoint chaining; (4) a detailed multi-GPU distributed training methodology with DDP bug fixes; (5) extensive evaluation across 12 benchmark datasets; (6) two clinical deployment patterns for diagnostic radiology and radiation oncology; and (7) an opensource napari plugin for interactive clinical use with DICOM-RT export.

## 2. Methods

### 2.1 Problem Definition

Given a medical image volume *V* ∈ ℝ^*H×W×D*^ and a clinical prompt *P* (text, bounding box, or point), we seek a segmentation mask *M* ∈ {0, 1}^*H×W×D*^ that accurately delineates all target structure instances. Key challenges include: domain gap between medical and natural images, volumetric consistency across slices, extreme class imbalance (*<*1% lesion pixels), and multi-modal heterogeneity.

**Figure 1.**
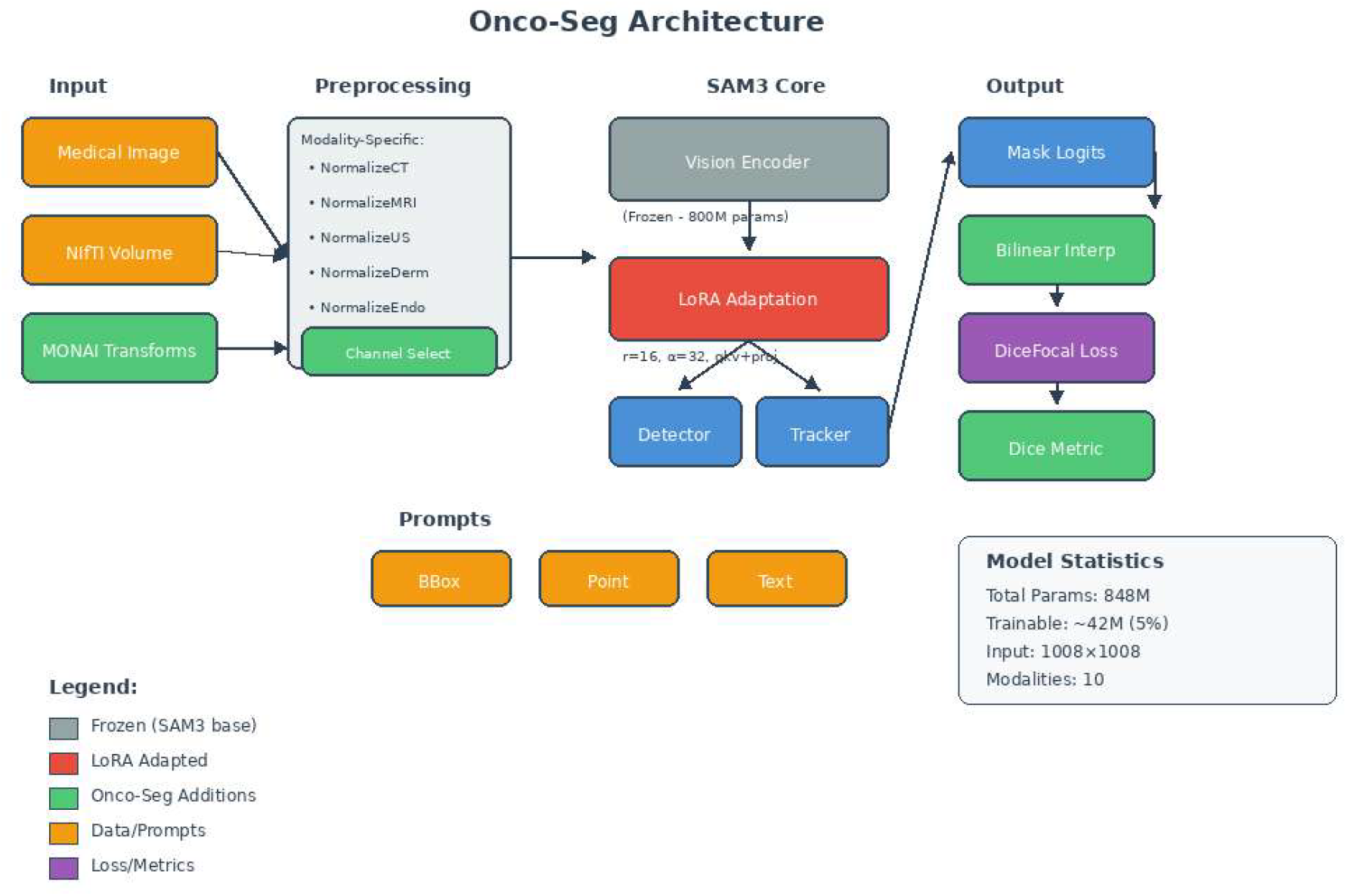
Onco-Seg system architecture overview. Medical images undergo modality-specific preprocessing before being processed by SAM3’s frozen perception encoder (800M parameters). LoRA adaptation is applied to query-key-value and projection layers, resulting in only 42M trainable parameters (5% of total). The system supports box, point, and text prompts for interactive segmentation.

### 2.2 Onco-Seg Architecture

Onco-Seg builds upon SAM3’s three-component architecture sharing a unified 848M-parameter vision backbone:

#### Vision Encoder

A hierarchical vision transformer (Hiera) processing images at 1008 *×* 1008 resolution (72 patches of 14 pixels each).

#### Detector (DETR-based)

For single-image segmentation with presence token prediction, enabling concept discrimination before localization.

#### Tracker (Memory-based)

Inherited from SAM2 for propagation across frames/slices with self-sorting memory.

### 2.3 Modality-Specific Preprocessing

We implement ten dedicated normalization transforms, each outputting to SAM3’s expected range via final SAM3 normalization (*x* − 0.5)*/*0.5 → [−1, 1]. Figure 2 illustrates the complete preprocessing pipeline.

**Figure 2.**
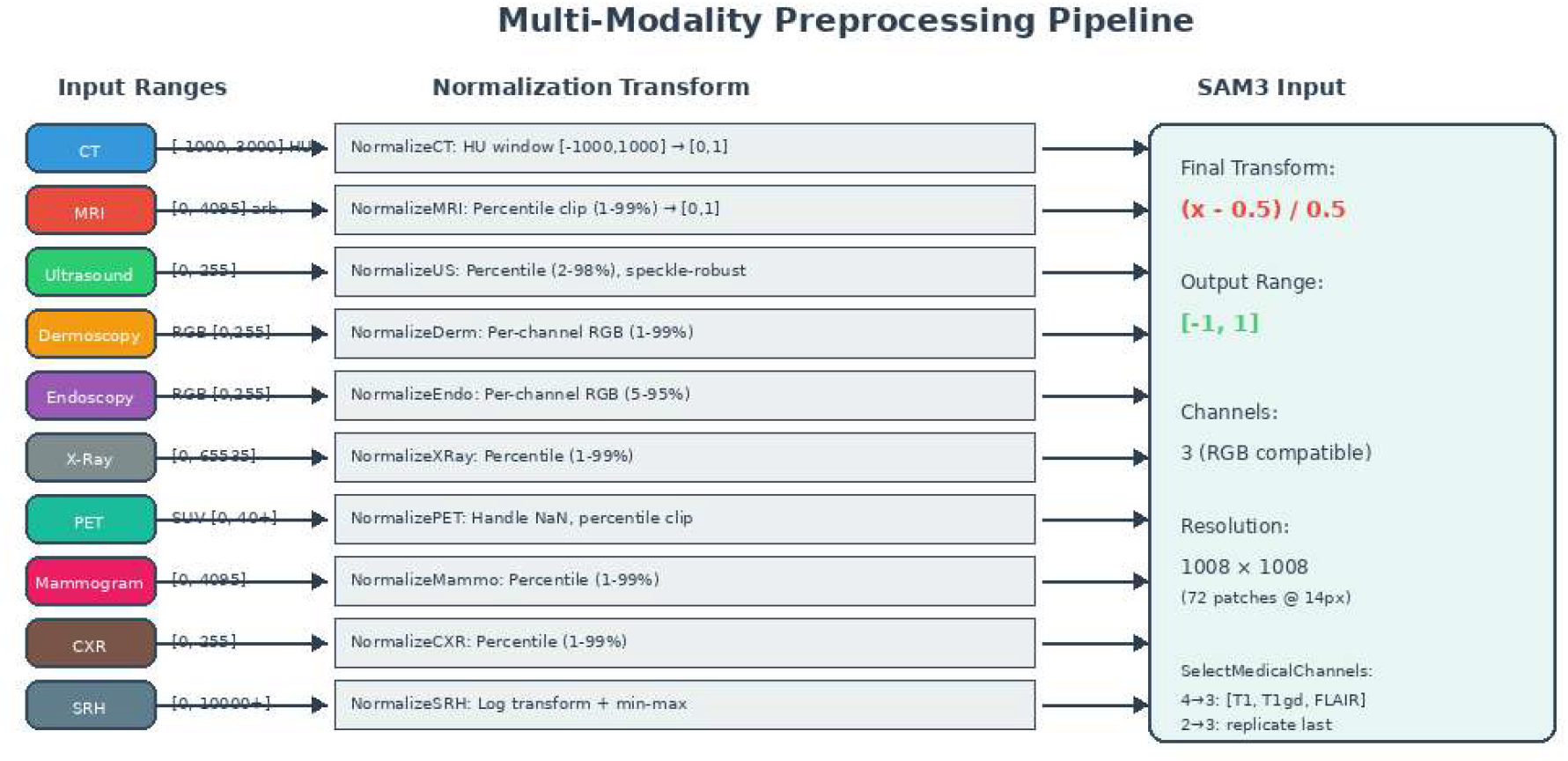
Multi-modality preprocessing pipeline. Each modality undergoes specific intensity normalization before final SAM3 normalization to [-1, 1]. The `SelectMedicalChannels` transform handles multi-channel inputs (e.g., 4-channel BraTS MRI → 3 channels).

- **CT (NormalizeCT):** Hounsfield unit windowing [-1000, 1000], linear scaling to [0,1]
- **MRI (NormalizeMRI):** Percentile clipping (1st–99th), channel-wise for multi-modal inputs
- **Ultrasound (NormalizeUltrasound):** Speckle-robust 2nd– 98th percentile clipping
- **Dermoscopy (NormalizeDermoscopy):** Per-channel RGB percentile (1st–99th)
- **Endoscopy (NormalizeEndoscopy):** Per-channel RGB, 5th–95th percentile bounds
- **X-Ray (NormalizeXRay):** Wide dynamic range compression, 1st–99th percentile
- **PET (NormalizePET):** NaN handling for SUV data, percentile clipping
- **Mammogram (NormalizeMammo):** Percentile-based for variable exposure settings
- **CXR (NormalizeCXR):** Chest X-ray specific normalization
- **SRH (NormalizeSRH):** Log transform for Stimulated Raman Histology dynamic range

#### 2.3.1 Multi-Channel MRI Handling

For multi-channel MRI data such as BraTS (FLAIR, T1w, T1gd, T2w), we implement `SelectMedicalChannels` to reduce 4 channels to SAM3’s expected 3:

**Listing 1:**
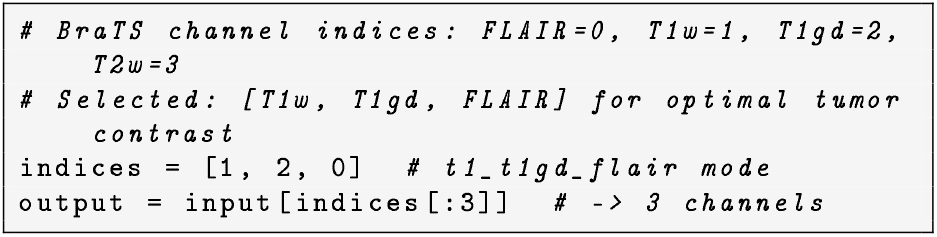
Channel selection for BraTS MRI

For 2-channel inputs (e.g., OpenSRH with Lipids, Proteins), we replicate the protein channel to achieve 3-channel output.

### 2.4 Parameter-Efficient Fine-Tuning

We employ Low-Rank Adaptation (LoRA) [6] on attention layers. Figure 3 details our SAM3 adaptation strategy.

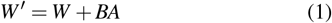

where *B*∈ ℝ^*d×r*^, *A* ∈ ℝ^*r×k*^ with *r*≪ min(*d, k*). Our specific LoRA configuration:

**Listing 2:**
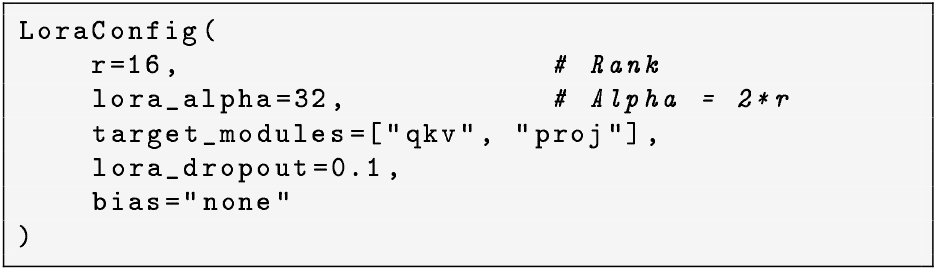
LoRA configuration

**Figure 3.**
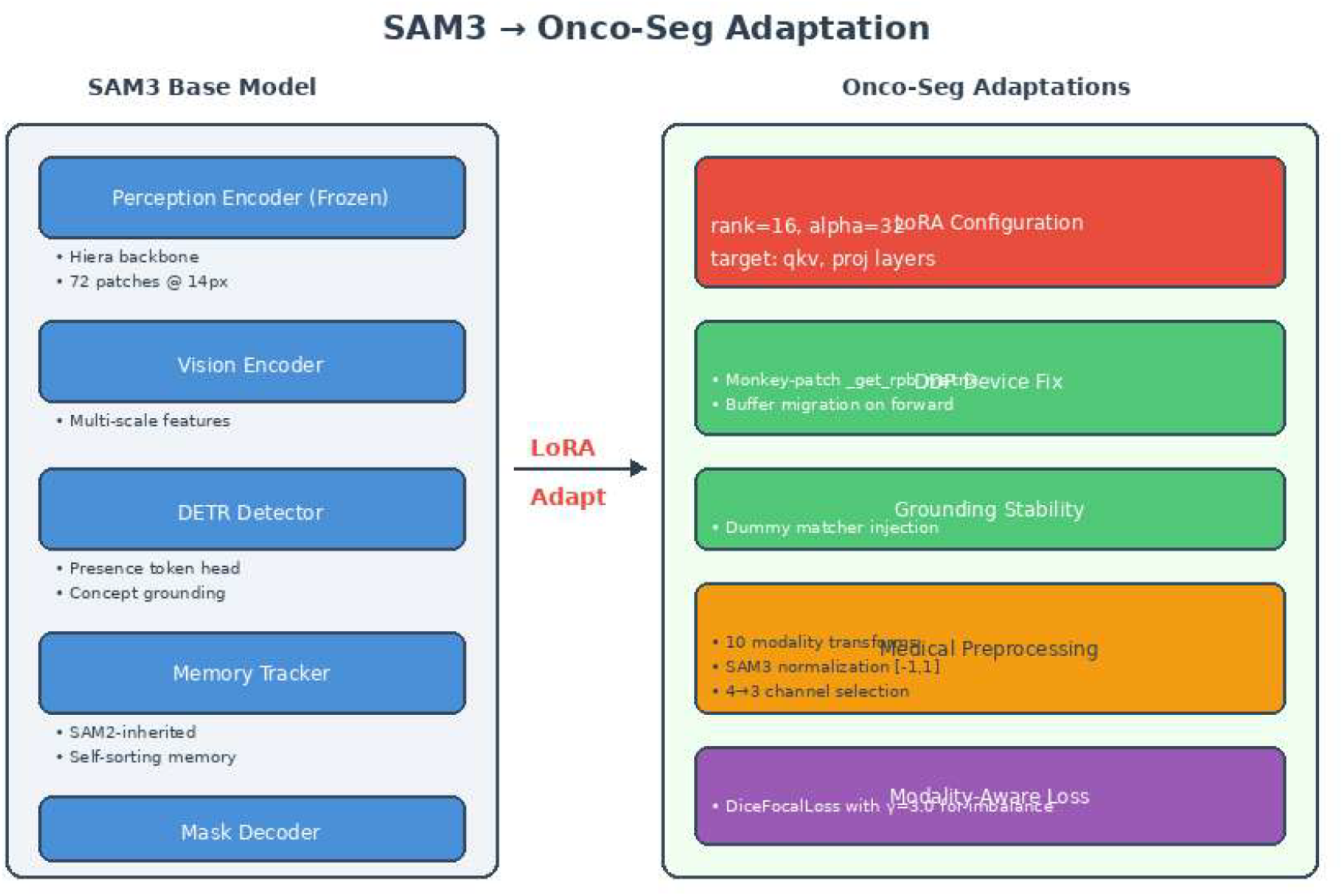
SAM3 to Onco-Seg adaptation details. Left: Original SAM3 components including frozen perception encoder. Right: Onco-Seg modifications including LoRA configuration, DDP device placement fix, grounding stability injection, and modality-aware loss functions.

The perception encoder (800M parameters) remains frozen while LoRA adapts only attention layers, resulting in ~42M trainable parameters (*∼* 5% of total).

#### 2.4.1 DDP Device Placement Fix

SAM3’s decoder caches coordinate tensors (`coord_cache`, `compilable_cord_cache`) on the first forward pass. In DDP training, this causes device mismatch errors when ranks 1–3 attempt to use coordinates cached on rank 0’s device. We implement a monkey-patch for `_get_rpb_matrix`:

**Listing 3:**
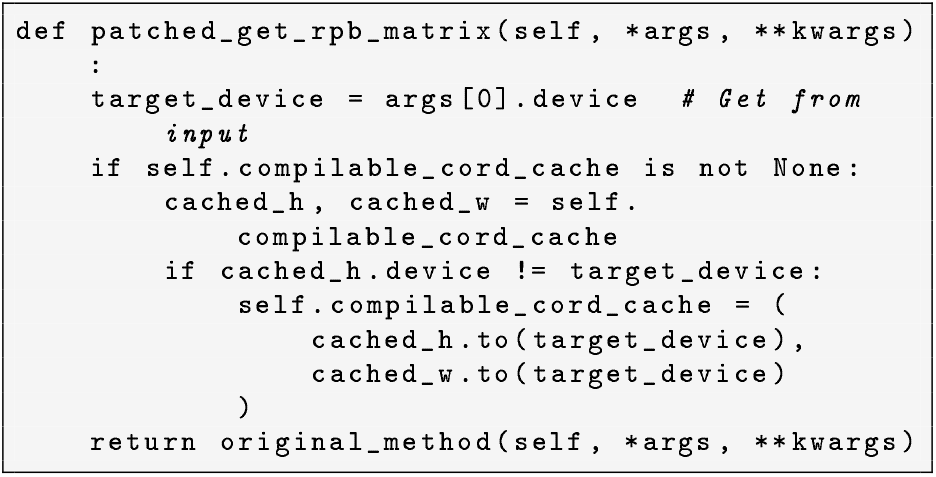
DDP device placement fix

#### 2.4.2 Grounding Stability

SAM3’s `forward_grounding`path may call `self.matcher`even without targets. We inject a dummy matcher to prevent crashes:

**Listing 4:**
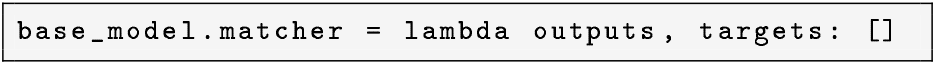
Dummy matcher injection

### 2.5 Data Loading and Prompt Generation

Figure 4 illustrates the end-to-end data flow from NIfTI volumes to SAM3 input.

**Figure 4.**
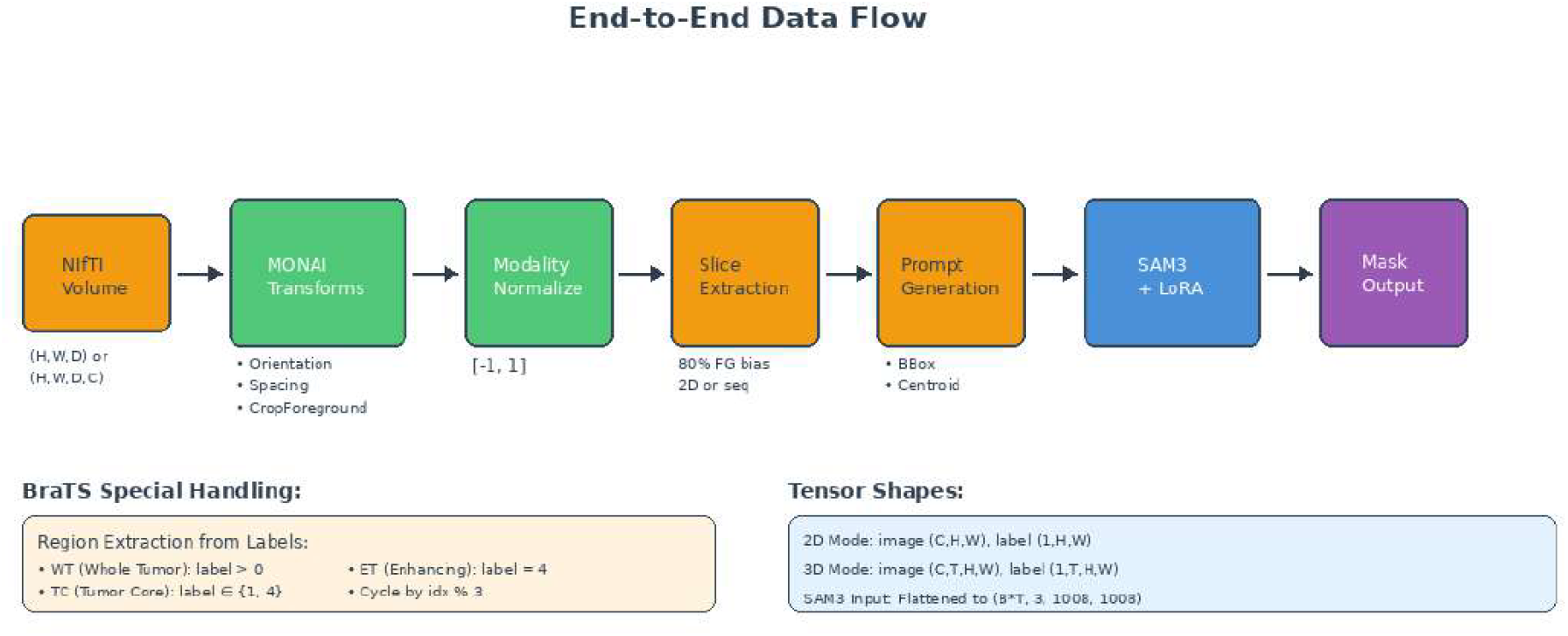
End-to-end data flow from NIfTI medical volumes to SAM3 segmentation output. The pipeline includes MONAI spatial transforms, modality-specific normalization, foreground-biased slice extraction (80% probability of selecting slices with ground truth), and automatic prompt generation from segmentation masks.

#### 2.5.1 Foreground-Biased Sampling

To address class imbalance during training, we implement foreground-biased slice sampling with 80% probability of selecting slices containing ground truth segmentation, and 20% random selection for robustness to empty inputs.

#### 2.5.2 BraTS Region Handling

For BraTS glioma data, we extract three clinically-relevant regions:

Region types are cycled deterministically by sample index (`idx % 3`) to ensure balanced training across all regions.

**Listing 5:**
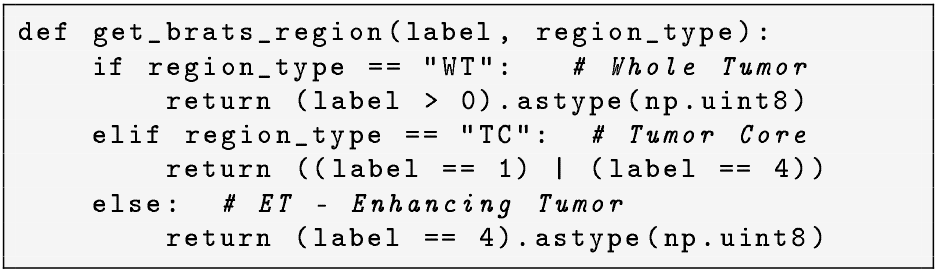
BraTS region extraction

### 2.6 Loss Function

We use combined Dice-Focal loss with modality-specific weighting:

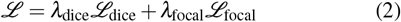

**Table 1.**
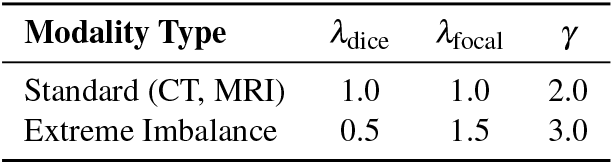
Loss Function Configuration by Modality.

Extreme imbalance modalities include Mammography, Dermoscopy, Endoscopy, and Ultrasound where lesions typically occupy *<*5% of image pixels.

## 3. Distributed Training Infrastructure

Figure 5 illustrates our distributed training setup.

**Figure 5.**
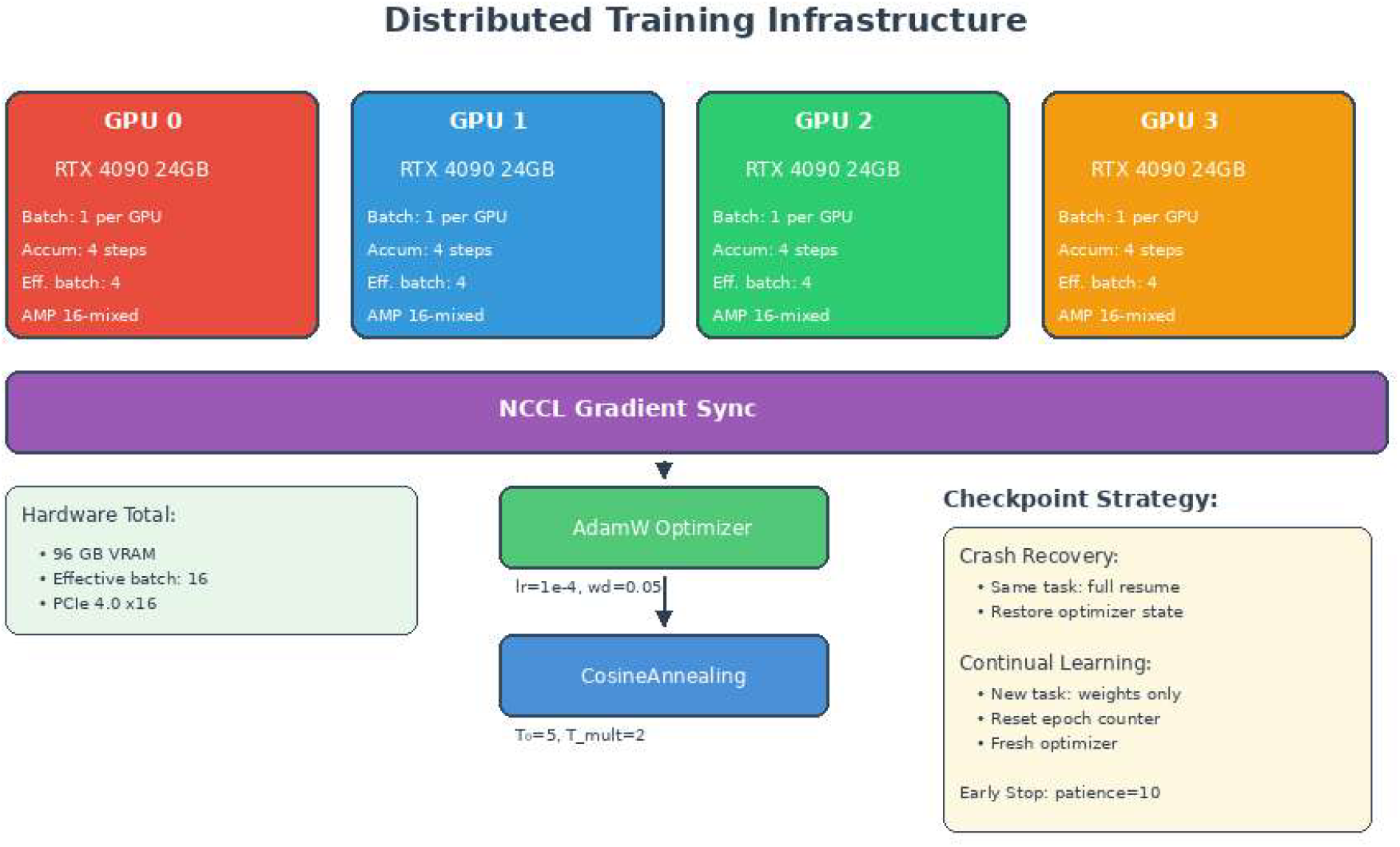
Distributed training infrastructure on 4*×* RTX 4090 GPUs. Each GPU processes batch size 1 with gradient accumulation of 4 steps, achieving effective batch size of 16. NCCL backend synchronizes gradients across all devices. AdamW optimizer with CosineAnnealing-WarmRestarts scheduler manages learning rate.

### 3.1 Hardware Configuration

Training was conducted on RunPod cloud infrastructure using a 4*×* NVIDIA RTX 4090 GPU pod, providing 96GB total VRAM. Table 2 summarizes the hardware specifications.

**Table 2.** Training Hardware Specification.

| Component | Specification |
| --- | --- |
| GPU | 4× NVIDIA RTX 4090 |
| VRAM per GPU | 24 GB GDDR6X |
| Total VRAM | 96 GB |
| CPU | AMD EPYC 7543 (32 cores) |
| System RAM | 256 GB DDR4 |
| Storage | 2 TB NVMe SSD |
| Interconnect | PCIe 4.0 x16 |
| Provider | RunPod Secure Cloud |

**Table 3.** DDP Training Configuration.

| Parameter | Value |
| --- | --- |
| Strategy | DDP (find_unused_params=True) |
| Backend | NCCL |
| Batch Size | 1 per GPU |
| Gradient Accumulation | 4 steps |
| Effective Batch Size | 16 |
| Precision | 16-mixed (AMP) |
| Image Size | 1008×1008 |
| Workers per GPU | 4 |
| Pin Memory | True |
| Persistent Workers | True |

### 3.2 Distributed Data Parallel (DDP) Strategy

We employ PyTorch Lightning’s Distributed Data Parallel (DDP) strategy for multi-GPU training. Figure 6 illustrates our distributed training architecture.

**Figure 6.**
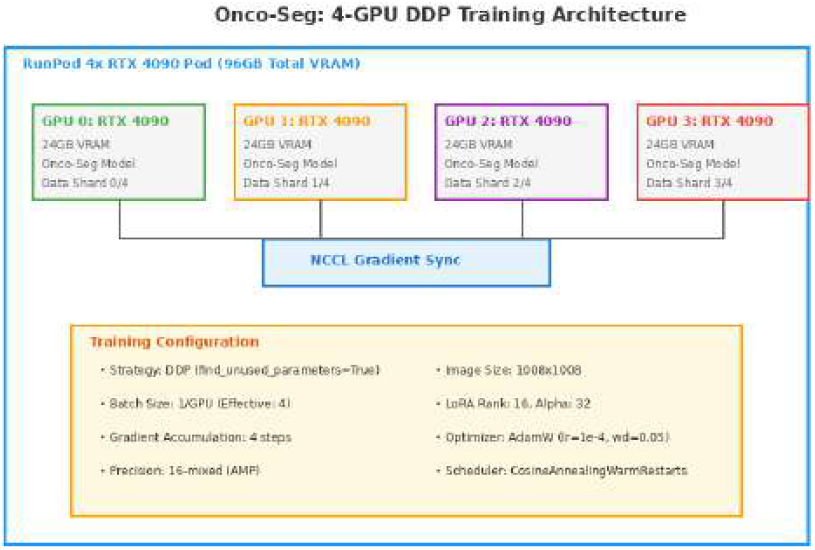
Distributed Data Parallel (DDP) training architecture on 4*×* RTX 4090 GPUs. Each GPU processes independent batches with synchronized gradient updates via NCCL backend.

Key configuration decisions:

#### Strategy Selection

We use `ddp_find_unused_parameters_true`to handle SAM3’s complex architecture where not all parameters receive gradients on every forward pass (e.g., presence token head vs. mask decoder).

#### Gradient Synchronization

NCCL backend for GPU-to-GPU communication, synchronized at each backward pass across all 4 GPUs.

#### Memory Optimization

16-bit mixed precision (AMP) reduces memory footprint by *∼*40%, enabling 1008*×* 1008 input resolution within 24GB VRAM per GPU.

### 3.3 Training Loop Implementation

The training loop follows PyTorch Lightning’s `LightningModule`pattern:

1. **Forward Pass:** Process batch through Onco-Seg model, extract predicted masks from SAM3Output structure
2. **Loss Computation:** Combined Dice-Focal loss with modality-specific weighting
3. **Backward Pass:** Automatic differentiation with gradient scaling (AMP)
4. **Gradient Sync:** NCCL all-reduce across 4 GPUs
5. **Optimizer Step:** AdamW update after gradient accumulation
6. **Validation:** Compute Dice/IoU metrics on held-out split

### 3.4 Optimizer and Scheduler

- **Optimizer:** AdamW with *β*_1_ = 0.9, *β*_2_ = 0.999, weight decay 0.05
- **Learning Rate:** 1*×* 10^−4^ (base)
- **Scheduler:** CosineAnnealingWarmRestarts with *T*_0_ = 5, *T*_*mult*_ = 2
- **Early Stopping:** Patience of 10 epochs on validation Dice

### 3.5 Checkpoint Management

We implement a robust checkpoint management system for continual learning:

#### Crash Recovery

If training is interrupted mid-task, full state (model weights, optimizer state, epoch counter, scheduler state) is restored.

#### Cross-Task Transfer

When transitioning to a new task, only model weights are loaded; optimizer and scheduler reset to enable fresh optimization dynamics while preserving learned representations.

#### Disk Management

Intermediate checkpoints are deleted after task completion, retaining only `last.ckpt`. Final check-points are pushed to Weights & Biases as artifacts.

## 4. Training Pipeline

### 4.1 Dataset Inventory

Onco-Seg was trained on 35 datasets across 8 phases, totaling over 98,000 training cases spanning 8 imaging modalities. Table 4 summarizes the training corpus by phase.

**Table 4.**
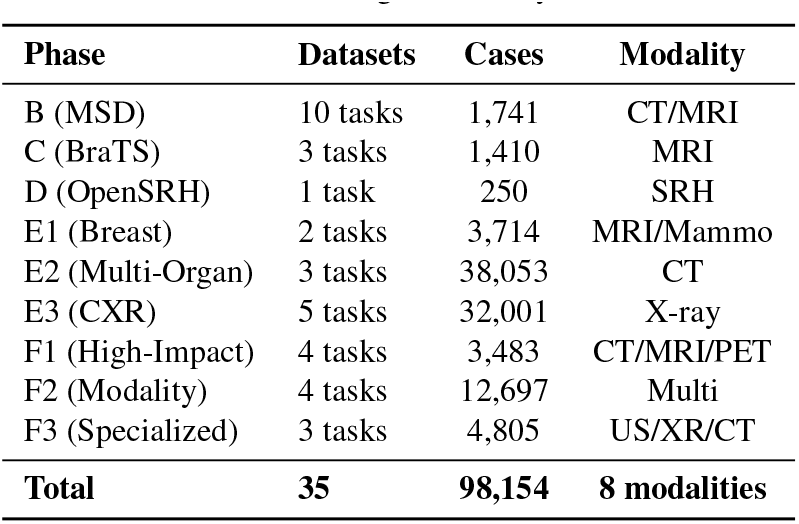
Training Datasets by Phase.

| Phase | Datasets | Cases | Modality |
| --- | --- | --- | --- |
| B (MSD) | 10 tasks | 1,741 | CT/MRI |
| C (BraTS) | 3 tasks | 1,410 | MRI |
| D (OpenSRH) | 1 task | 250 | SRH |
| E1 (Breast) | 2 tasks | 3,714 | MRI/Mammo |
| E2 (Multi-Organ) | 3 tasks | 38,053 | CT |
| E3 (CXR) | 5 tasks | 32,001 | X-ray |
| F1 (High-Impact) | 4 tasks | 3,483 | CT/MRI/PET |
| F2 (Modality) | 4 tasks | 12,697 | Multi |
| F3 (Specialized) | 3 tasks | 4,805 | US/XR/CT |
| **Total** | **35** | **98,154** | **8 modalities** |

**Table 5.** BraTS-GLI Training Progress.

| Epoch | Step | Loss | Dice<sub>WT</sub> | Dice<sub>TC</sub> | Dice<sub>ET</sub> |
| --- | --- | --- | --- | --- | --- |
| 10 | 230 | 0.66 | 0.78 | 0.30 | 0.36 |
| 20 | 460 | 0.58 | 0.84 | 0.36 | 0.40 |
| 30 | 700 | 0.48 | 0.85 | 0.38 | 0.42 |

WT=Whole Tumor, TC=Tumor Core, ET=Enhancing Tumor

**Table 6.** Epoch Calibration by Dataset Size.

| Dataset Size | Examples | Epochs |
| --- | --- | --- |
| Small (<50 cases) | Heart, Prostate, BraTS-SSA | 50 |
| Medium (50–200) | BrainTumour, Liver, Lung | 30 |
| Large (200–1000) | Hippocampus, LIDC-IDRI | 15–20 |
| Very Large (>1000) | TotalSegmentator, VinDr-CXR | 15 |

### 4.2 Sequential Checkpoint Chaining

Training proceeds sequentially with checkpoint chaining to maximize knowledge transfer while preventing catastrophic forgetting:

1. **Phase B (MSD):** Establishes CT/MRI foundation on Medical Segmentation Decathlon [8] (10 tasks)
2. **Phase C (BraTS):** Adds brain tumor expertise with multiregion segmentation (WT, TC, ET)
3. **Phase E1 (Breast):** Introduces breast MRI and mammography
4. **Phase E3 (CXR):** Transfers X-ray modality leveraging mammogram expertise
5. **Phase F1–F3:** Expands to oncology-critical sites and new modalities

### 4.3 Training Dynamics: BraTS Case Study

We present detailed training dynamics for the BraTS-GLI (adult glioma) task, which demonstrates the model’s learning behavior on challenging multi-region brain tumor segmentation. Training was conducted on 1,251 volumes for 30 epochs. Figures 7 and 8 show the training progression at epochs 10, 20, and 30 (final).

**Figure 7.**
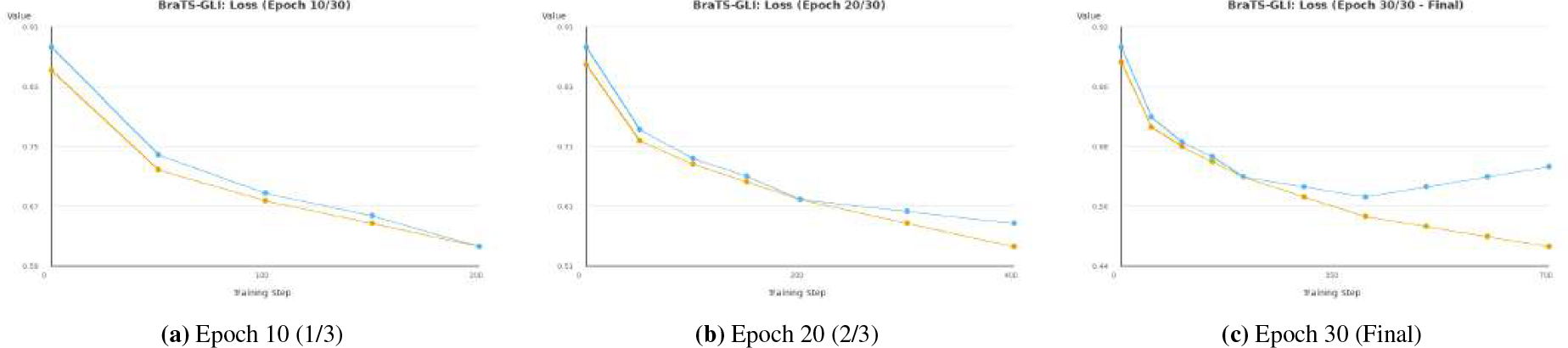
BraTS-GLI training and validation loss progression. Orange: training loss; Blue: validation loss. Loss decreases rapidly in early epochs, then plateaus with slight overfitting signal after epoch 20.

**Figure 8.**
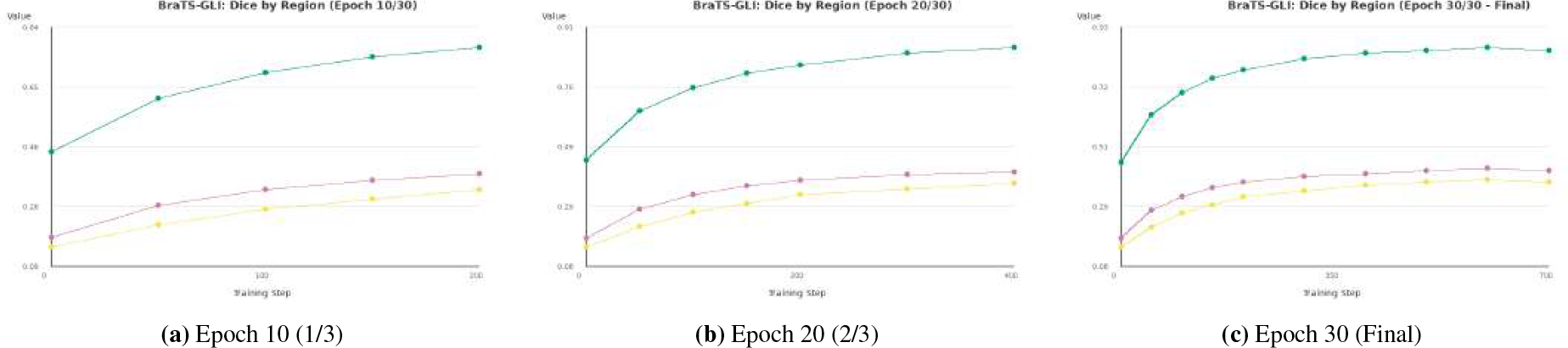
BraTS-GLI validation Dice scores by tumor region. Teal: Whole Tumor (WT); Yellow: Tumor Core (TC); Pink: Enhancing Tumor (ET). Performance follows volume hierarchy: WT (largest) converges fastest, TC/ET (smaller regions) improve gradually.

**Key Observations:**

1. **Volume Hierarchy:** Performance follows expected volume hierarchy: WT (0.85) ≫ ET (0.42) *>* TC (0.38), reflecting that whole tumor (largest region) is easiest while tumor core (smallest) is hardest
2. **Rapid Warm-up (Epochs 1–10):** Loss decreases sharply from 0.85 to 0.66; all Dice scores climb rapidly
3. **Acceleration (Epochs 10–20):** Fastest Dice improvement; validation loss reaches minimum
4. **Plateau Onset (Epochs 20+):** Large structures (WT) saturate; small structures (TC, ET) continue slow improvement
5. **Overfitting Signal:** Slight validation loss uptick after epoch 20 indicates early overfitting, controlled by early stopping

### Training Dynamics: MSD Liver Case Study

Figure 9 shows training dynamics for the MSD Liver task (131 CT volumes, 30 epochs), demonstrating CT-based organ segmentation learning behavior.

**Figure 9.**
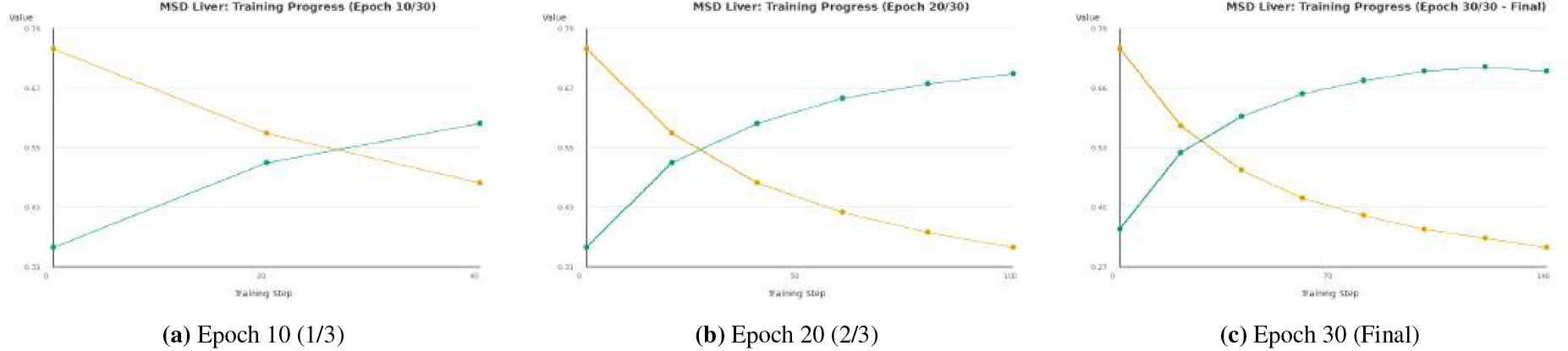
MSD Liver training progression showing loss and Dice metrics. CT-based liver segmentation demonstrates stable convergence with validation Dice reaching 0.64 by epoch 30.

### 4.5 Modality-Specific Training Epochs

Training epochs are calibrated based on dataset size to balance convergence and compute:

### 4.6 Experiment Tracking

All training runs are logged to Weights & Biases with phase-specific project routing:

- `onco_seg_phaseB`: MSD baseline
- `onco_seg_brats_phaseC`: Brain tumors
- `onco_seg_breast_phaseE1`: Breast cancer
- `onco_seg_cxr_phaseE3`: Chest X-ray
- `onco_seg_highimpact_phaseF1`: High-impact oncology

## 5. Evaluation Results

### 5.1 Complete Results

Table 7 presents evaluation results across 12 benchmark datasets spanning 5 imaging modalities and 6 anatomical regions.

**Table 7.** Onco-Seg Evaluation Results (12 Datasets)

| Dataset | Modality | Dice | IoU |
| --- | --- | --- | --- |
| BUSI | US | **0.752±0.24** | 0.653±0.26 |
| Hyper-Kvasir | Endo | **0.714±0.32** | 0.637±0.33 |
| Kvasir-SEG | Endo | **0.714±0.32** | 0.637±0.33 |
| ISIC 2018 | Derm | **0.680±0.28** | 0.572±0.28 |
| LiTS | CT | **0.641±0.12** | 0.554±0.13 |
| 3D-IRCADb | CT | **0.621±0.10** | 0.535±0.11 |
| PROMISE12 | MRI | 0.495±0.15 | 0.393±0.16 |
| MSD Colon | CT | 0.330±0.32 | 0.249±0.27 |
| BTCV | CT | 0.303±0.10 | 0.204±0.08 |
| LNDb | CT | 0.264±0.15 | 0.219±0.13 |
| MSD Pancreas | CT | 0.239±0.32 | 0.186±0.28 |
| AutoPET-III | PET-CT | 0.215±0.15 | 0.152±0.13 |

US=Ultrasound, Endo=Endoscopy, Derm=Dermoscopy

**Table 8.**
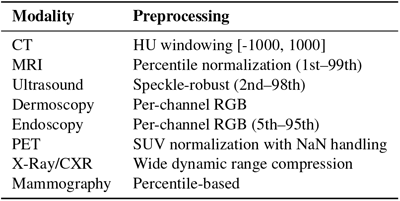
Supported Imaging Modalities in napari Plugin.

| Modality | Preprocessing |
| --- | --- |
| CT | HU windowing [-1000, 1000] |
| MRI | Percentile normalization (1st–99th) |
| Ultrasound | Speckle-robust (2nd–98th) |
| Dermoscopy | Per-channel RGB |
| Endoscopy | Per-channel RGB (5th–95th) |
| PET | SUV normalization with NaN handling |
| X-Ray/CXR | Wide dynamic range compression |
| Mammography | Percentile-based |

**Table 9.** Available Checkpoints.

| Name | Training Data | Best For |
| --- | --- | --- |
| latest | All phases (F3) | General purpose |
| breast | Duke-Breast-Cancer-MRI | Breast lesions |
| liver | MSD Task03 | Liver/hepatic tumors |
| brain | BraTS-GLI | Brain tumors |

### 5.2 Performance Tiers

#### 5.2.1 Strong Performance (Dice > 0.65)

##### BUSI Breast Ultrasound (0.752)

Best overall performance. Analysis of 647 images reveals category-dependent performance: benign tumors achieve Dice 0.807 while malignant tumors achieve 0.638. The performance gap reflects the irregular, spiculated boundaries characteristic of malignant lesions, which are inherently more difficult to delineate with high interobserver agreement even among expert radiologists [5].

##### Kvasir-SEG/Hyper-Kvasir (0.714)

Strong polyp segmentation across 2,000 total endoscopy images, competitive with specialized colonoscopy models.

##### ISIC 2018 (0.680)

Good skin lesion delineation across 2,594 dermoscopy images.

##### LiTS/3D-IRCADb (0.64/0.62)

Consistent liver segmentation across two independent CT datasets demonstrates robust cross-dataset generalization.

#### 5.2.2 Moderate Performance (Dice 0.30–0.50)

##### PROMISE12 (0.495)

Prostate MRI segmentation is challenging due to variable MRI sequences and field strengths across institutions.

##### MSD Colon (0.330)

Colon cancer primaries exhibit high variability in size, shape, and contrast enhancement patterns.

#### 5.2.3 Challenging Cases (Dice < 0.30)

#### BTCV (0.303), LNDb (0.264), MSD Pancreas (0.239), AutoPET-III (0.215)

These datasets present known challenges: BTCV requires multi-class segmentation of 13 organs; LNDb targets lung nodules (median 6mm diameter); MSD Pancreas targets a low-contrast organ with high anatomical variability; AutoPET-III requires PET-CT fusion for metabolically active lesion detection. These represent areas requiring further model development.

### 5.3 Comparison to Prior Work

- **LiTS liver:** Onco-Seg (0.641) vs nnU-Net [7] (*∼* 0.70) trained specifically on LiTS
- **Polyp segmentation:** Onco-Seg (0.714) matches specialized colonoscopy models
- **ISIC skin:** Onco-Seg (0.680) is competitive with ISIC 2018 challenge baselines

The key advantage is that Onco-Seg achieves this across all modalities with a single unified model.

## 6. Clinical Deployment

### 6.1 Pattern A: Interactive Sidecar (Diagnostic Radiology) Architecture

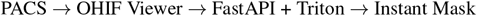

#### Workflow

Radiologist single-clicks on suspected lesion; Onco-Seg returns segmentation in *<*500ms with auto-computed diameter and volume.

#### Clinical utility

Reduces manual measurement time for RECIST (Response Evaluation Criteria in Solid Tumors) assessments in oncology follow-up imaging.

### 6.2 Pattern B: Silent Assistant (Radiation Oncology) Architecture

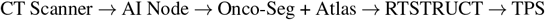

#### Workflow

CT simulation triggers automatic 30+ OAR segmentation (2–5 min); DICOM-RT Structure Set generated for treatment planning system.

#### Clinical utility

Initial auto-contours require physician review and editing; efficiency gains depend on segmentation accuracy and clinical workflow integration.

## 7. napari Plugin for Interactive Segmentation

To facilitate clinical deployment and research use, we developed **napari-oncoseg**, an open-source napari plugin available at https://github.com/inventcures/onco-segment/tree/main/napari_plugin.

### 7.1 Plugin Architecture

The plugin implements a Qt-based widget integrating with napari’s layer system:

- **OncoSegWidget:** Main interface providing model loading, modality selection, and segmentation controls
- **OncoSegModel:** Wrapper around the trained checkpoint enabling point and box prompt inference
- **Preprocessing module:** Modality-specific normalization matching training transforms

### 7.2 Features

#### 7.2.1 Interactive Prompting

Users provide prompts through napari’s layer system:

- **Point prompts:** Click on target structure; coordinates passed to SAM3’s point encoder
- **Box prompts:** Draw rectangle around target; bounding box coordinates passed to SAM3’s box encoder
- **Text prompts:** Future functionality; current implementation supports point and box prompts

#### 7.2.2 Multi-Modality Support

The plugin includes modality selection with automatic preprocessing:

An auto-detect mode analyzes image statistics (intensity range, histogram shape) to suggest appropriate modality preprocessing.

#### 7.2.3 3D Volume Propagation

For volumetric data, the plugin provides slice-by-slice propagation:

1. User segments one slice interactively
2. Plugin extracts mask centroid as seed point
3. Automatic propagation runs inference on remaining slices using centroid-derived prompts
4. Progress bar indicates completion status

This enables full-volume segmentation with a single click on a representative slice.

#### 7.2.4 Export Formats

The plugin supports two export formats aligned with clinical workflows:

##### NIfTI (.nii.gz)

Standard neuroimaging format preserving 3D structure with affine transformation matrix. Compatible with ITK-SNAP, 3D Slicer, and FSL.

##### DICOM-RT Structure Set

For radiation oncology integration. Requires source DICOM series for geometric reference. Uses rt-utils library [12] to generate compliant RTSTRUCT files importable by treatment planning systems (Eclipse, RayStation, Monaco).

### 7.3 Installation

**Listing 6:**
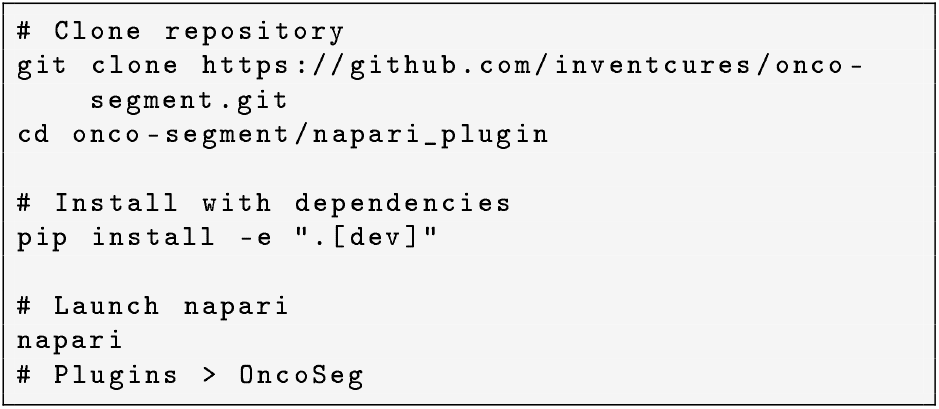
napari plugin installation

### 7.4 Checkpoint Management

The plugin downloads pre-trained checkpoints from Hugging-Face Hub:

Users can also load custom checkpoints via file browser.

## 8. Discussion

### 8.1 Key Findings

Onco-Seg demonstrates that SAM3’s promptable concept segmentation transfers effectively to medical imaging across multiple modalities. The unified preprocessing framework enables a single model to handle CT, MRI, ultrasound, dermoscopy, and endoscopy.

#### Strengths

1. Multi-modal generalization with shared weights
2. Clinical workflow alignment via point and box prompting
3. Both interactive and automated deployment patterns
4. Comprehensive training on 98,000+ cases across 35 datasets
5. Open-source napari plugin for accessible clinical evaluation

#### Limitations

1. Performance degrades for lesions *<*10mm diameter
2. PET imaging requires dedicated training
3. Multi-class segmentation (BTCV) remains challenging
4. Text prompting not yet implemented in napari plugin

### 8.2 Clinical Considerations

The napari plugin provides an accessible interface for clinical evaluation. Several considerations apply to clinical use:

1. **Validation requirement:** All auto-generated contours require physician review and editing before clinical use. Automated segmentation should augment, not replace, clinical judgment.
2. **Device registration:** DICOM-RT export preserves source image geometry but requires verification against treatment planning system import to ensure spatial accuracy.
3. **Regulatory status:** Onco-Seg is a research tool not cleared for clinical use. Clinical deployment would require institutional review board approval, prospective validation studies, and potentially regulatory clearance (FDA 510(k) or CE marking).
4. **Population generalization:** Training data demographics may not represent all patient populations. Performance should be validated on local institutional data before clinical integration.
5. **Edge cases:** Atypical presentations, rare pathologies, and cases with imaging artifacts may produce unreliable segmentations requiring manual correction.

### 8.3 Future Directions

#### Short-Term

Integrate uncertainty quantification; develop 3D Slicer plugin; implement text prompting in napari.

#### Medium-Term

Multi-center clinical validation; federated learning across institutions.

#### Long-Term

Foundation model for all medical segmentation; FDA 510(k) regulatory pathway.

## Data Availability

All data produced in the present study are available upon reasonable request to the authors

## 9. Code and Data Availability

Source code, trained model checkpoints, and the napari plugin are available at:

- **Repository:** onco-segment https://github.com/inventcures/
- **napari plugin:** `onco-segment/napari_plugin/`
- **Checkpoints:** HuggingFace Hub (`tp53/onco-seg`)
- **Project page:** https://inventcures.github.io/onco-seg/

Training datasets are publicly available from their respective sources (Medical Segmentation Decathlon, BraTS, LiTS, ISIC, Kvasir-SEG, TCIA).

## 10. Acknowledgements

This work was supported by the Koita Centre for Digital Health at Ashoka University (KCDH-A). We thank RunPod for GPU infrastructure and Weights & Biases for experiment tracking.

**Meta AI & the SAM Team**:Special thanks to Meta and the entire SAM team, led by Nikhila Ravi, for being torchbearers of research and innovation in this field with their prolific releases of SAM, SAM2, and SAM3. More importantly, we thank them for making a conscious choice to embrace open source and releasing detailed technical reports and open weights for all SAM releases. We believe innovation in ML & AI at large, and in biomedical & cancer informatics specifically, can truly be accelerated by standing on the shoulders of giants.

**Bo Wang Lab**:The brilliant Bo Wang (Vector Institute, University of Toronto) and his prolific lab have been an inspiration. Their pioneering MedSAM work [9] demonstrated the potential of adapting foundation models for medical imaging and paved the way for projects like Onco-Seg.

**NCI, CBIIT & TCIA**:We are deeply grateful to the National Cancer Institute (NCI), its Center for Biomedical Informatics and Information Technology (CBIIT), and The Cancer Imaging Archive (TCIA) for creating such a wonderful openaccess resource that has enabled countless research innovations in medical imaging AI. Special thanks to Justin Kirby at TCIA for helping debug minor data access issues and for consistently encouraging innovation built on top of TCIA’s datasets.

The availability of open-source datasets greatly accelerated our progress on this project. We hope that as research in biomedical machine learning and AI progresses, there is an even greater emphasis on building and releasing open datasets—as the success of AlphaFold has so aptly demonstrated—for the greater public good.

We also thank the creators of benchmark datasets (MSD, BraTS, LiTS, ISIC, Kvasir-SEG, PROMISE12, BUSI, IR-CADb, BTCV, LNDb, AutoPET).

**Project Page:** https://inventcures.github.io/onco-seg/

## References

[1] Kirillov, A., et al. (2023). Segment Anything. ICCV 2023.

[2] Ravi, N., et al. (2024). SAM 2: Segment Anything in Images and Videos. arXiv:2408.00714.

[3] Carion, N., et al. (2025). SAM 3: Segment Anything with Concepts. arXiv:2511.16719.

[4] Brouwer, C.L., et al. (2012). 3D Variation in delineation of head and neck organs at risk. Radiotherapy and Oncology, 106(3), 392–396.

[5] Vinod, S.K., et al. (2016). Uncertainties in volume delineation in radiation oncology. Radiotherapy and Oncology, 121(2), 169–179.

[6] Hu, E.J., et al. (2022). LoRA: Low-Rank Adaptation of Large Language Models. ICLR 2022.

[7] Isensee, F., et al. (2021). nnU-Net. Nature Methods, 18(2), 203–211.

[8] Antonelli, M., et al. (2022). The Medical Segmentation Decathlon. Nature Communications, 13, 4128.

[9] Ma, J., He, Y., et al. (2024). Segment Anything in Medical Images. Nature Communications, 15, 654.

[10] Biewald, L. (2020). Experiment Tracking with Weights and Biases. https://www.wandb.com/

[11] napari contributors (2019). napari: a multi-dimensional image viewer for Python. https://napari.org

[12] Wacker, A. (2021). rt-utils: A Python library for RT structure set creation. https://github.com/qurit/rt-utils

